# Synthetic Echocardiograms from Diffusion Models in Rare Cardiovascular Disease

**DOI:** 10.64898/2026.09.14.26363024

**Authors:** Pengfei Lyu, Ricardo Henao, Lydia Coulter Kwee, Fred Zhangzhi Peng, Melissa Hurdle, Sreekanth Vemulapalli, Svati H. Shah, Michel Georges Khouri, Anru Zhang

## Abstract

The limited availability of imaging data for uncommon cardiovascular phenotypes constrains the development of robust imaging models. We evaluated whether class-conditional diffusion models can generate synthetic transthoracic echocardiograms that improve downstream cardiac imaging tasks. The primary application was cardiac amyloidosis detection in a Duke University cohort using a two-step classifier, with external analyses using EchoNet-Dynamic for image-fidelity assessment and EchoNet-LVH for wall-thickness phenotype classification. The Duke cohort was partitioned at the patient-encounter level into 70% training, 15% validation, and 15% test sets; generators were trained only on the training partition, augmentation levels were selected using validation AUROC, and final evaluation used held-out real test data. In the Duke all-view two-step analysis, adding synthetic images increased AUROC from 0.883 to 0.924, with an AUROC difference of 0.041 (95% CI, 0.013–0.069); in EchoNet-LVH, AUROC increased from 0.832 to 0.864, with an AUROC difference of 0.032 (95% CI, 0.021–0.044). Expert review found that synthetic images were sometimes difficult to identify as synthetic, but rated them lower for diagnostic adequacy. These findings suggest that diffusion-generated echocardiograms may provide a practical approach to augmenting limited training data for selected cardiac imaging tasks and motivate further evaluation across clinical settings.

## 1 Introduction

The limited availability of imaging examples for uncommon cardiovascular phenotypes can constrain the development of robust machine-learning models. Transthoracic echocardiography (TTE) is widely used to assess myocardial structure, chamber geometry, ventricular function, and other cardiac phenotypes, and routine clinical acquisition provides a substantial source of imaging data for automated cardiovascular phenotyping. Machine-learning and deep-learning models have shown promise across echocardiographic tasks, including view classification, ejection-fraction estimation, and disease screening [1–4]. More broadly, machine learning has become increasingly important in medicine and in supporting rare-disease diagnosis [5, 6]. Despite the overall abundance of clinical imaging data, clinically important diseases and phenotypes may remain sparsely or unevenly represented in routine imaging archives, limiting the number and diversity of labeled examples available for model development.

This scarcity often manifests as class imbalance in echocardiographic datasets, particularly for rare diseases, severe functional abnormalities, uncommon structural phenotypes, and challenging differential-diagnosis groups. Standard approaches to class imbalance include random over- or under-sampling, cost-sensitive learning, threshold adjustment, and synthetic minority oversampling methods such as SMOTE and its variants [7–15]. Resampling can increase the representation of existing minority-class examples but cannot introduce new phenotypic variation. This limitation may be particularly important for high-dimensional echocardiographic data, in which image appearance depends on anatomy, imaging view, acquisition conditions, and image quality. Generative augmentation offers an alternative approach by producing additional examples from a learned data distribution, although its utility should ultimately be assessed on held-out real data rather than inferred from visual plausibility alone [16–18].

Cardiac amyloidosis (CA) is an infiltrative cardiomyopathy whose echocardiographic findings can overlap with hypertensive heart disease and other causes of left ventricular hypertrophy (LVH), particularly with respect to increased ventricular wall thickness [19]. CA detection therefore provides a clinically relevant setting in which limited disease prevalence produces class imbalance while phenotypic overlap further complicates classification.

Denoising diffusion probabilistic models (DDPMs) [20, 21] and related score-based models [22] have emerged as powerful approaches for image synthesis. Relative to generative adversarial networks [23, 24], diffusion models can provide stable training and diverse sampling behavior [25]. Their utility for echocardiographic synthesis and downstream data augmentation across heterogeneous disease and phenotype-classification tasks, however, remains incompletely characterized. Related diffusion-based work in medical imaging and synthetic electronic health records provides a foundation for evaluating both synthetic-image fidelity and downstream task performance [17, 18, 26].

We therefore evaluated diffusion-generated echocardiograms using CA detection in a Duke University cohort as the primary disease-classification application, a setting characterized by both class imbalance and substantial phenotypic overlap with LVH. We additionally evaluated two external echocardiography datasets: EchoNet-Dynamic for LVEF-conditioned generation and image-fidelity assessment, and EchoNet-LVH for wall-thickness phenotype classification. Across these settings, we assessed image fidelity, expert evaluation, retention of task-relevant information using synthetic-only training, and the effect of synthetic augmentation on classification performance using held-out real data, with ordinary minority-class resampling as a comparator. An overview of the study design is shown in Figure 1.

**Figure 1:**
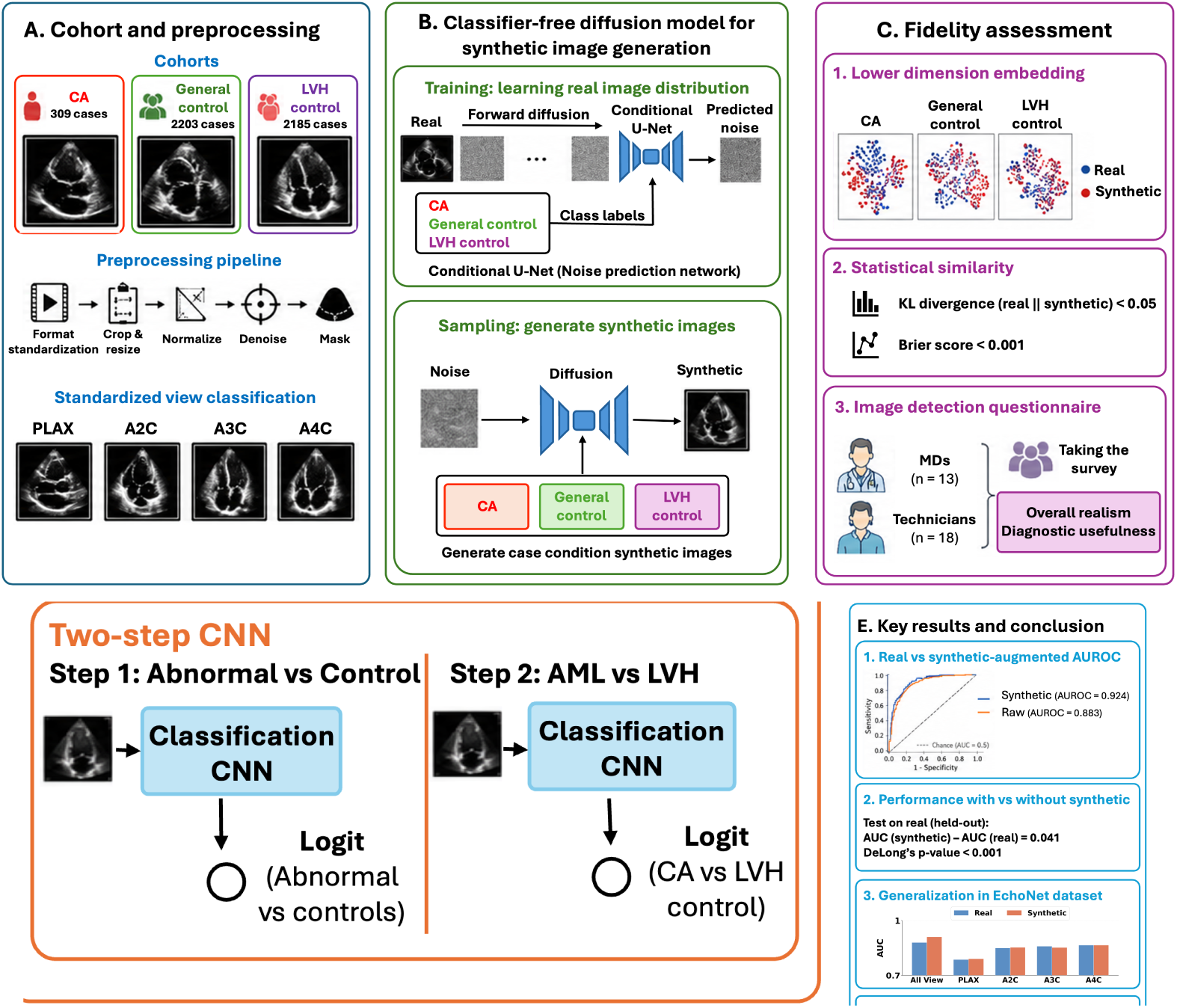
Study overview. (**a**) Cohort composition and preprocessing pipeline. The cohort includes cardiac amyloidosis (CA) cases, general controls without evidence of cardiac hypertrophy, and left ventricular hypertrophy (LVH) controls. The preprocessing pipeline includes format standardization, resizing, quality control, registration, and masking. The images are classified into parasternal long-axis view (PLAX), apical two-chamber view (A2C), apical three-chamber view (A3C), and apical four-chamber view (A4C). (**b**) Class-conditional diffusion models for task-specific synthetic transthoracic echocardiogram generation, with learned class embeddings and 1,000 forward diffusion steps during training; sampling uses 500 reverse denoising steps. (**c**) Fidelity assessment comprising two-dimensional visualization, pooled grayscale-intensity comparison, discriminator analysis, and expert evaluation. (**d**) Downstream classification using a convolutional neural network (CNN) under real-only, synthetic-only, bootstrap, and real-plus-synthetic training strategies. For the retained primary Duke CA application, the two-step model first distinguishes abnormal cases from general controls and then distinguishes CA from LVH. (**e**) External task evaluations: EchoNet-Dynamic provides external LVEF-conditioned generation and fidelity evaluation, whereas EchoNet-LVH provides external wall-thickness phenotype classification. The panels summarize the visual, distributional, expert, and downstream classification analyses. Changes in performance varied by task, and the figure does not imply a uniform benefit or universal generalizability.

## 2 Methods

### 2.1 Study Design and Evaluation Tasks

We evaluated a diffusion-based framework for generating synthetic echocardiograms across complementary internal and external tasks. The Duke CA all-view two-step classification task was the primary analysis: the comparison was the change in patient-encounter-level AUROC between real-only and real-plus-synthetic training. Secondary analyses included synthetic-only versus real-only training, the EchoNet-LVH wall-thickness task, and comparison with bootstrap resampling. External fidelity analyses were exploratory or descriptive and are reported in the Supplementary Information. All final classification evaluations used held-out real echocardiograms.

### 2.2 Datasets

#### 2.2.1 Duke Cardiac Amyloidosis Cohort

Transthoracic echocardiogram (TTE) studies performed for clinical indications were retrospectively obtained from the Duke University Health System echocardiography archive after eligible encounters were assigned to one of three prespecified phenotype groups: cardiac amyloidosis (CA), left ventricular hypertrophy (LVH) controls, or general controls without LVH. CA cases were encounters from patients with a documented clinical diagnosis of CA before or at the time of the eligible echocardiogram; labels were adjudicated by expert review by a cardiologist with expertise in CA. LVH controls were encounters from patients with evidence of LVH and no known CA diagnosis. General controls were encounters from patients without known CA and without evidence of cardiac hypertrophy. LVH was included as a particularly challenging control group because it shares key structural features with CA, most notably increased myocardial wall thickness, which can produce similar echocardiographic appearances and make differential diagnosis difficult without additional clinical or histopathological information. General controls were included to represent the broader population without cardiac hypertrophy, enabling evaluation of model performance under both difficult and easier discrimination settings and to mirror clinical diagnostic scenarios.

Within these phenotype definitions and acquisition windows, all available eligible TTE encounters with DICOM imaging were retained before image-level preprocessing; no random sampling from a larger eligible pool and no case-control matching were performed. The CA cohort included studies acquired between June 2004 and March 2024, while general control and LVH control studies were collected between January 2015 and December 2023. The full image-level Duke dataset contained 2,346 CA images, 5,831 general control images, and 6,013 LVH-control images after DICOM and image-quality preprocessing. These image counts are distinct from the actual patient-encounter counts used for partitioning and final evaluation. The cohort comprised 309 CA encounters, 2,203 general-control encounters, and 2,185 LVH-control encounters, for a total of 4,697 patient encounters. Each study corresponded to a unique patient encounter. The primary split was stratified by encounter rather than by unique patient: all images from the same patient encounter were assigned to one partition, but the current split file does not establish that a patient with multiple encounters could not appear in more than one partition. Final area under the receiver operating characteristic (AUROC) and area under the precision-recall curve (AUPRC) were evaluated at the patient-encounter level after image-level predicted probabilities were averaged within encounter. The patient-encounter partition counts are summarized in Table 1. Because the split was encounter-level, patient-level independence was not established.

**Table 1:** Duke cohort partition counts by patient encounter.

| Group | Training | Validation | Test | Total |
| --- | --- | --- | --- | --- |
| CA | 216 | 46 | 47 | 309 |
| General controls | 1,542 | 330 | 331 | 2,203 |
| LVH controls | 1,529 | 328 | 328 | 2,185 |
| Total | 3,287 | 704 | 706 | 4,697 |

#### 2.2.2 EchoNet-Dynamic

EchoNet-Dynamic [2] was used for LVEF-conditioned image generation and external fidelity evaluation. EchoNet-Dynamic consists technically of apical four-chamber (A4C) echocardiographic videos and was not treated as a multi-view dataset. The videos are labeled by left ventricular ejection fraction (LVEF); because the dataset does not include CA or LVH labels, it was not used for CA detection. Instead, frames were grouped into LVEF categories and used to compare real and synthetic image distributions. The analysis included three LVEF classes: normal (LVEF ≥ 55%, n = 6,961), mild dysfunction (40% ≤ LVEF < 55%, n = 805), and severe dysfunction (LVEF < 40%, n = 1,246).

#### 2.2.3 EchoNet-LVH

EchoNet-LVH [27] was used for external wall-thickness phenotype classification because it provides a standardized benchmark with a single echocardiographic view, predefined training, validation, and test partitions, and expert-derived cardiac measurements. EchoNet-LVH contains 12,000 deidentified parasternal long-axis echocardiographic videos acquired during routine clinical care at Stanford Medicine, with expert-derived measurements of cardiac structure including interventricular septal thickness, left ventricular internal dimension, and left ventricular posterior wall thickness. After excluding 349 videos with missing interventricular septal thickness at end-diastole (IVSd) or left ventricular posterior wall thickness at end-diastole (LVPWd), 11,651 videos were retained. For each video, maximum diastolic wall thickness was defined as the larger of IVSd and LVPWd. The minority phenotype threshold was determined exclusively from the training set as the 90th percentile of this value (1.3274), and the same threshold was then applied unchanged to the validation and test sets. This yielded 10,399 majority-group videos and 1,252 minority-group videos overall: the training set contained 9,152 majority and 1,019 minority videos, the validation set contained 1,037 majority and 104 minority videos, and the test set contained 210 majority and 129 minority videos.

The EchoNet-LVH phenotype label was derived from end-diastolic IVSd and LVPWd measurements, whereas the classifier input was the predefined first frame of each video. The first frame was not selected to correspond to end diastole; therefore, this analysis evaluates prediction of a measurement-derived wall-thickness phenotype from a standardized first-frame representation rather than classification from the measurement frame itself.

### 2.3 Image and Video Preprocessing

An automated preprocessing pipeline was implemented to curate raw DICOM files. Non-standard acquisitions and color Doppler images were excluded based on DICOM metadata fields, including SOP Class UID and ultrasound color flow indicators, to ensure consistency in imaging modality and view representation for downstream model development. For each retained echocardiographic video, only the first frame was extracted and retained as the real-image representation. No additional frames from the same video were used as separate training examples. The diffusion model was trained on these single-frame images using the training partition and generated single synthetic images during inference. The CNN classifier used the same first-frame modality, where the input contains the single-frame real images or the synthetic images generated via the diffusion model. Validation and test evaluation used real first-frame images only. When more than one image was available for a patient encounter, image-level predicted probabilities were averaged before calculating encounter-level performance metrics. The first-frame rule was predefined and consistently applied; it was not intended to identify a particular cardiac phase or the most informative image.

To screen for corrupted or non-diagnostic frames, an automated quality-control pipeline was implemented at the image level. For each frame, eight quantitative features were extracted as image-quality screening variables: (i) sector coverage, measuring the proportion of the image area occupied by the ultrasound imaging wedge; (ii) non-black pixel ratio, defined as the fraction of pixels with positive grayscale intensity, where zero-valued pixels represented black background outside the ultrasound sector or other blank image regions rather than a physiological measurement; (iii–iv) intensity-based summary statistics, specifically the mean and standard deviation of pixel values across the frame; (v) Shannon entropy, capturing image texture and intensity variation; (vi– vii) higher-order distributional moments, comprising skewness and kurtosis of the pixel intensity distribution; and (viii) edge density, summarizing the amount of boundary information present. Feature values were standardized to zero mean and unit variance across the dataset. Frames for which at least one quality-control feature had an absolute standardized value greater than 3 (|Z | > 3) were flagged for review. This threshold was used as a high-sensitivity screening rule rather than as an automatic exclusion criterion. Final exclusion required manual confirmation that the frame was unusable for image-based modeling, such as a blank or corrupted frame, absent or severely truncated ultrasound sector, non-echocardiographic content, residual color Doppler or annotation artifacts that obscured the cardiac field, or image quality too poor to identify cardiac structures. Frames flagged by the quantitative screen but not meeting these visual exclusion criteria were retained.

All retained images underwent a standardized preprocessing workflow before model development. This pipeline included spatial resizing to 112 × 112 pixels and center-based cropping to ensure uniform image dimensions. To promote anatomical consistency across samples, images were partially registered by detecting sector boundaries, fitting a circular geometric model to the ultrasound field, and applying translation, rotation, and affine transformations to align all sectors to a shared reference coordinate system. Finally, a sector-based masking procedure was applied to isolate the ultrasound imaging region while removing extraneous elements such as text overlays, electrocardiographic traces, and device-generated annotations.

### 2.4 Class-Conditional Diffusion Model

For each generation experiment, a task-specific class-conditional denoising diffusion probabilistic model (DDPM) [20, 21] was trained using the relevant task label y. In the Duke CA cohort, y represented the three phenotype groups: CA, LVH controls, and general controls. In EchoNet-Dynamic, y represented the three LVEF categories: normal, mild dysfunction, and severe dysfunction. In EchoNet-LVH, y represented the binary wall-thickness phenotype. Thus, separate dataset-specific generators were trained for the Duke CA, EchoNet-Dynamic, and EchoNet-LVH tasks. The U-Net architecture is illustrated in Figure 2. Generators were trained only on the relevant training partition, and validation and test images were excluded from generator training.

**Figure 2:**
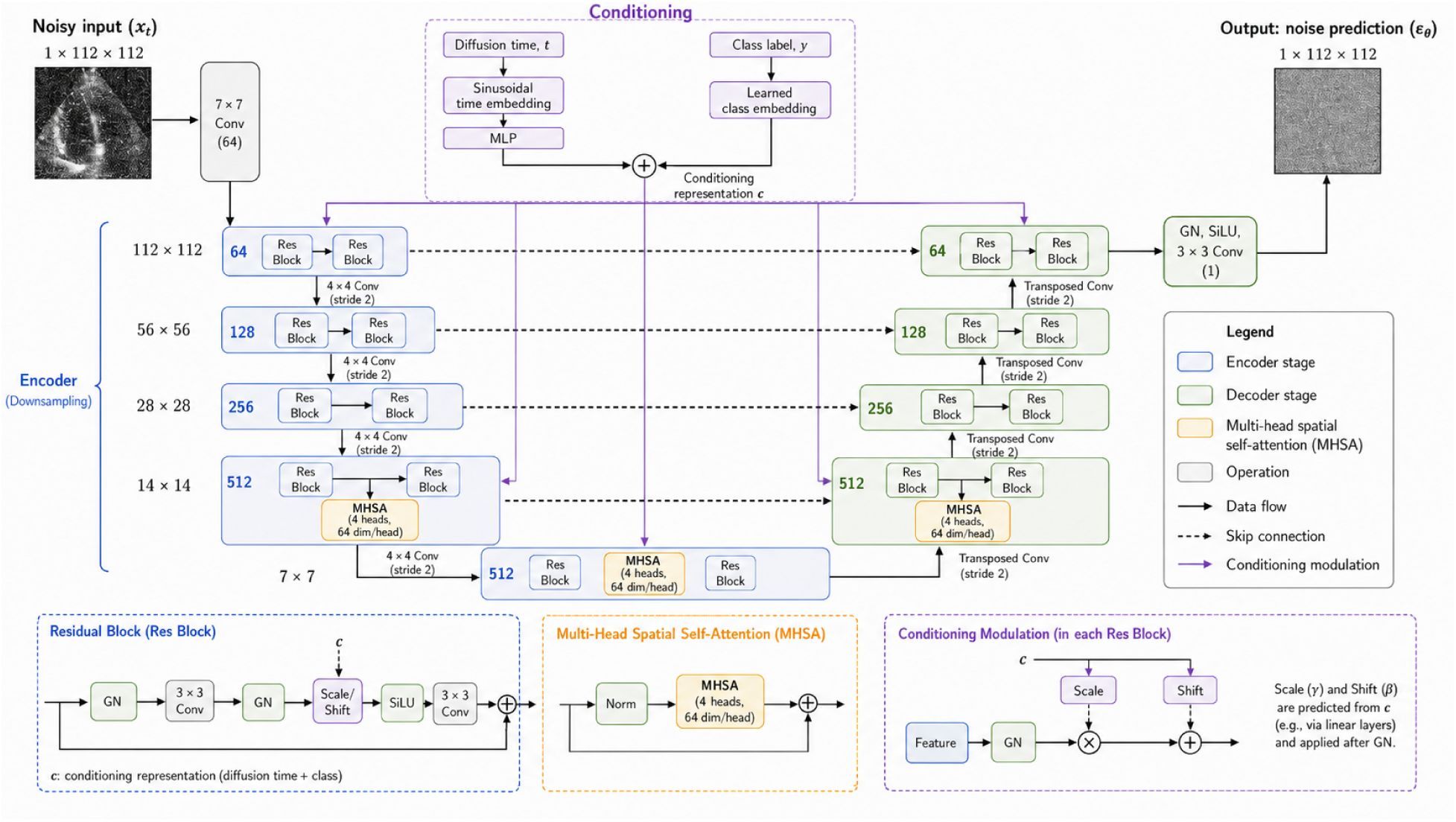
Class-conditional U-Net for diffusion-based echocardiographic image generation. The network receives a noisy single-channel image x_t_ with dimensions 1 × 112 × 112 and predicts the added noise ε_θ_ at diffusion step *t*. The diffusion-time input is transformed by a sinusoidal time embedding and multilayer perceptron, while the task label *y* is transformed by a learned class embedding. These representations are added to form the conditioning representation c, which modulates each residual block through scale-and-shift conditioning. The U-Net contains encoder and decoder stages with 64, 128, 256, and 512 channels, skip connections, and multi-head spatial self-attention at the deepest encoder and decoder resolutions and in the bottleneck. The forward diffusion process used 1,000 steps during training, and inference used 500 reverse DDIM denoising steps to generate a synthetic echocardiographic image under the specified class condition.

The denoising network was implemented as a two-dimensional class-conditional U-Net in PyTorch. It received single-channel 112 × 112 grayscale echocardiographic images as input. An initial 7 × 7 convolution mapped the input to 64 feature channels. The encoder and decoder comprised four resolution stages with 64, 128, 256, and 512 feature channels, respectively. Each stage contained two residual blocks, and skip connections linked corresponding encoder and decoder stages. Each residual block used two 3 × 3 convolutions, group normalization, SiLU activation, and modulation by the diffusion-time and class-conditioning representation. Downsampling used 4 × 4 stride-2 convolutions, and upsampling used transposed convolutions. Multi-head spatial self-attention was applied at the deepest encoder and decoder resolutions and in the bottleneck [28], using four attention heads with 64 dimensions per head. The final block consisted of group normalization, SiLU activation, and a 3 × 3 convolution that returned a single-channel noise prediction with the same spatial dimensions as the input.

Let *x*_0_ denote the original echocardiographic image, *x_t_* the noisy image after diffusion step t, and y the class-conditioning label. We write 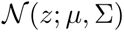 for a Gaussian distribution over *z* with mean *µ* and covariance matrix Σ, and **I** for the identity covariance matrix. In the forward process, a real image *x*_0_ was corrupted over *T* = 1,000 timesteps using the Markov chain 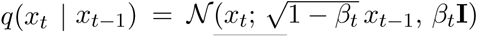, where {*β_t_*} followed a sigmoid schedule. Equivalently, 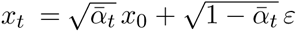, with 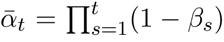 and 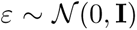. The reverse transition was written as 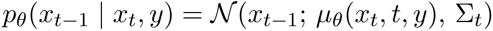, where the U-Net *ε*_θ_ predicted the noise added at step t, and *µ*_θ_ and the schedule-derived covariance Σ*_t_* defined the reverse transition.

Diffusion-step conditioning used a 16-dimensional sinusoidal time encoding followed by a multilayer perceptron that produced a 256-dimensional conditioning vector. The task label entered the U-Net through a learned class embedding of the same dimension, which was added to the time representation. Classifier-free guidance was enabled by randomly dropping the class label for 10% of training examples and replacing it with a learned null-class embedding [29]. At sampling time, the conditional and null-label noise estimates were combined as 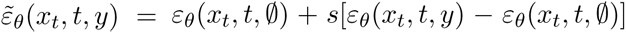, where s ≥ 1 controls the contribution of the conditional estimate. Models were trained with the Adam optimizer using mini-batches of 16 images and a learning rate of 8 × 10**^-^**^5^, with exponential moving averages of the model parameters.

The forward diffusion process therefore used *T* = 1,000 steps during training. At inference, synthetic images were generated from Gaussian noise using 500 reverse DDIM denoising steps [30] conditioned on the target class label.

### 2.5 Downstream Echocardiographic Classification Tasks

We evaluated synthetic data in downstream tasks involving disease status, ventricular function, and myocardial wall thickness. Unless otherwise specified, the same classifier architecture and model-selection procedure were used across real-only, synthetic-only, bootstrap, and real-plus-synthetic experiments. For the Duke CA task, CNN classifiers used the first extracted frame from each echocardiographic video as a still-image input. The retained CA analysis used the same CNN backbone and first-frame input modality for both stages of the two-step framework and the corresponding downstream experiments. The CNN comprised four 3 × 3 convolution–ReLU–max-pooling blocks with 32, 64, 128, and 256 channels, followed by adaptive average pooling, dropout (p = 0.3), and a two-class fully connected classifier head. EchoNet-LVH used this same CNN backbone in a single-stage binary classifier for the wall-thickness phenotype. Models were optimized using the Adam optimizer with a cosine learning rate schedule. During training, standard data augmentation techniques were applied, including random horizontal flipping, small-angle rotations (up to ±10**^◦^**), and random adjustments to brightness and contrast, to improve model robustness and reduce overfitting.

#### 2.5.1 Synthetic Data Generation and Augmentation

We considered three training paradigms: (i) real-only training, establishing a real-only reference benchmark; (ii) synthetic-only training; and (iii) real-plus-synthetic training, in which synthetic CA echocardiograms were added only to the training partition. Candidate synthetic-sample counts were evaluated from 0 to 2,000 in increments of 100. For each candidate count, the classifier was trained using the training partition and evaluated on the validation partition. The candidate yielding the highest validation AUROC was selected, and the selected configuration was evaluated once on the held-out test partition. Each candidate was fit once under the experiment run settings, and ties were resolved in favor of the smaller synthetic-sample count. The validation selection curve was retained as an analysis output, but task-specific selected counts are not included in the current manuscript. The test partition was not used to select the number of synthetic samples, the classifier, the training epoch, or hyperparameters.

#### 2.5.2 Cardiac Amyloidosis Classification

The retained Duke CA classifier used a two-step framework to reflect the clinical difficulty of separating CA from LVH. Stage 1 separated abnormal cases (CA and LVH combined) from general controls, and Stage 2 distinguished CA from LVH among abnormal cases. For an image x, the final continuous CA score was the product of the Stage 1 abnormal probability and the Stage 2 conditional CA probability, P (CA | x) = P (abnormal | x)P (CA | abnormal, x). These probabilities were composed at the image level and then averaged within encounter before calculation of encounter-level performance. This two-step design separates detection of structural abnormality from differentiation of CA and LVH. Synthetic CA images were added to the training data for both stages. Because Stage 1 contained 1,745 abnormal encounters (216 CA and 1,529 LVH) versus 1,542 general controls in the training partition, CA-targeted augmentation in Stage 1 should not be interpreted as conventional minority-class augmentation; Stage 2 was the CA-versus-LVH minority-class setting.

### 2.6 Statistical Analysis

Model performance was evaluated on an encounter-level holdout partition that was defined before model training and kept unchanged across the real-only, synthetic-only, and real-plus-synthetic experiments. The Duke cohort was split at the patient-encounter level into 70% training, 15% validation, and 15% test partitions, with all images from a given patient encounter assigned to the same partition to prevent leakage; the validation partition was used for model selection, and the test partition was reserved for final evaluation. All confidence intervals (CIs) were calculated as 95% CIs. Because multiple images could be available from the same patient encounter, image-level predicted probabilities were first averaged within encounter, and all performance estimates and uncertainty intervals were then calculated at the patient-encounter level. For individual AUROC estimates, 95% CIs were estimated using the nonparametric DeLong method [31]. For comparisons between real-only and real-plus-synthetic models evaluated on the same held-out encounters, 95% CIs for the paired AUROC differences were calculated using the paired DeLong comparison variance.

Primary performance metrics were computed at the patient-encounter level, matching the unit used for data splitting and avoiding the independence assumption that would be violated by treating multiple images from the same encounter as independent observations. Image-level model outputs were used only as intermediate predictions: when multiple images were available for the same patient encounter, their predicted probabilities were averaged to obtain a single patient-level score before calculating AUROC and AUPRC. AUPRC was available as a point estimate for the secondary TRTR/TSTR diagnostic but is not reported for the primary augmentation table because a verified primary-task AUPRC output was not available in the current analysis record. For the bootstrap baseline, real minority-class training samples were resampled with replacement. The number of added bootstrap samples matched the number of synthetic minority-class training samples used in the corresponding diffusion-augmentation experiment. Majority-class training data were left unchanged, and a new bootstrap sample was generated for each classifier training run. All remaining classifier architecture, optimization, model-selection, and other training settings were held constant.

To systematically assess the utility of synthetic data as a training resource, we implemented three evaluation paradigms following the framework of Tian et al. [26]. **Train-on-Real-Test-on-Real (TRTR):** models are trained on real transthoracic echocardiograms and evaluated on held-out real echocardiograms, establishing the real-only reference benchmark. **Train-on-Synthetic-Test-on-Real (TSTR):** models are trained exclusively on synthetic echocardiograms and evaluated on held-out real echocardiograms, assessing whether synthetic data retain task-relevant information. **Train-on-Real-and-Synthetic-Test-on-Real (TRSTR):** real training data are augmented with synthetic echocardiograms and evaluated on held-out real echocardiograms, assessing downstream augmentation utility. These paradigms were applied where permitted by the available labels and study design. Synthetic echocardiograms were used only in training partitions; validation and test partitions contained only real echocardiograms.

### 2.7 Synthetic Image Fidelity Evaluation

Synthetic image fidelity was evaluated using complementary visual, distributional, and discriminative analyses. The t-distributed stochastic neighbor embedding (t-SNE) and Uniform Manifold Approximation and Projection (UMAP) plots were treated as qualitative two-dimensional visualizations rather than formal tests of distributional equality. These plots were used to visualize relationships between synthetic and real transthoracic echocardiograms in a reduced two-dimensional representation.

To compare global grayscale intensity distributions, images were loaded using Pillow and converted to 8-bit grayscale. For each image set, pixel-intensity counts were accumulated across images using a 256-bin NumPy histogram over the range [0, 256). An additive constant of 10**^-^**^12^ was applied to every bin before normalization. Kullback–Leibler (KL) and Jensen–Shannon divergences were then calculated using custom Python functions. Images were not resized or equally weighted before pooling; thus, each image contributed in proportion to its number of pixels. The analysis pooled all eligible EchoNet-Dynamic real images in `EchoNet-Dynamic/First Frames` and all corresponding generated images in `EchoNet-Dynamic/Synthetic Images/Generated Diffusion`, without stratification by LVEF category. No external benchmark or prespecified comparator was available, so these quantities were used only as descriptive diagnostics. Both directions of KL divergence were calculated because KL is asymmetric, while Jensen–Shannon divergence was used as a symmetric and smoothed comparison. Jensen–Shannon distance, the square root of Jensen– Shannon divergence, and RMSE were retained as descriptive quantities; all values are reported in Supplementary Table S1 and were not treated as independent fidelity analyses. Let *p_b_* and *q_b_* denote the normalized real and synthetic probabilities for grayscale-intensity bin *b*, where *b* = 1, . . ., 256. To provide a directly interpretable measure on the probability scale, distribution-level RMSE was computed between the normalized 256-bin grayscale-intensity histograms:

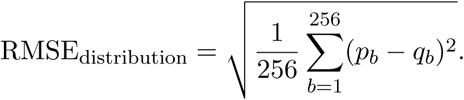

This metric was not derived from classifier probabilities and did not involve model training or data partitioning. We did not report the corresponding mean squared difference separately because it contains the same information as RMSE after a square-root transformation.

We also trained a real-versus-synthetic discriminator using the Duke CA dataset to quantify separability between real and synthetic echocardiograms. The discriminator used the same CNN backbone described above, with single-channel 112 × 112 images as input and a binary output distinguishing real from synthetic images. The discriminator data were partitioned at the patient level into 70% training, 15% validation, and 15% test sets, with observations associated with the same patient restricted to a single partition. The validation set was used to select the checkpoint with the highest encounter-level AUROC; training was stopped after a maximum of 20 epochs or after 5 consecutive validation epochs without improvement. The final discriminator was evaluated once on the held-out test set. In this analysis, an AUROC of 0.5 would indicate no discrimination between the two image sources. The observed test AUROC was interpreted together with its confidence interval, partition design, and class composition.

### 2.8 Expert Survey Design

To evaluate the perceptual and clinical fidelity of synthetic CA transthoracic echocardiograms from a human perspective, we conducted a blinded online survey distributed to physicians and cardiac ultrasound technicians at Duke University Health System. The survey presented 20 single echocardiograms — 10 real and 10 synthetic CA images, presented in randomized order — without disclosing which images were synthetic. For each image, respondents rated: (i) echocardiographic view, (ii) identifiable anatomical structures, (iii) brightness, (iv) ROI presentation, (v) diagnostic adequacy, (vi) confidence in interpretation, (vii) visual realism on an ordinal agreement scale, and (viii) perceived image source, categorized as definitely or probably real, unsure, or probably or definitely synthetic. Thus, the visual-realism item assessed whether the image looked natural, whereas the source-classification item asked respondents to infer whether the image came from a real or synthetic source. A paired comparison task was also included, asking respondents to judge which of two simultaneously presented images appeared more realistic. Participants saw the paired images only as the first and second image; responses were recoded after unblinding as preferences for the real image, the synthetic image, or neither image. The survey was initiated by 31 respondents, including 13 physicians and 18 cardiac ultrasound technicians. Completed surveys were available from 17 respondents, including 6 physicians and 11 cardiac ultrasound technicians. The core image-level questions yielded 199 real-image evaluations and 193 synthetic-image evaluations after item-wise exclusion of missing responses. Percentages and summary statistics used the number of non-missing responses for each item as the denominator. Response entropy was computed at the image level using base-2 logarithms, 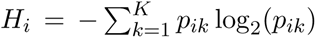, where *p_ik_* is the proportion of responses in category k for image i. Entropy was normalized by the maximum possible entropy, log_2_ (*K*), where *K* denotes the number of response categories for that item: 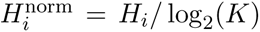. Normalized entropy ranged from 0 to 1 and was averaged within real and synthetic image groups. Because each evaluator rated multiple images, responses were not treated as independent for confirmatory inference. Survey analyses were primarily descriptive. Paired-image comparisons were summarized as supportive evidence of perceptual ambiguity, and no confirmatory hypothesis test was performed. The survey was approved under the same institutional protocol as the imaging study.

## 3 Results

Results are presented in three parts: synthetic-image fidelity assessment, synthetic-only task-retention diagnostics, and real-data augmentation experiments. The Duke all-view two-step CA classification task was the primary endpoint. EchoNet-LVH wall-thickness classification and bootstrap comparison were secondary analyses, while fidelity analyses were exploratory or descriptive.

### 3.1 Fidelity of Generated Echocardiograms

We evaluated generated echocardiograms using complementary visual, histogram-based, discriminative, and expert-review analyses. Figure 3 shows t-SNE and UMAP visualizations for EchoNet-Dynamic, a public external dataset used here for LVEF-conditioned fidelity assessment. Across the three LVEF classes defined in the Methods (normal, mild dysfunction, and severe dysfunction), real and synthetic samples show visual overlap in the two-dimensional visualizations. We included both t-SNE and UMAP because they emphasize different aspects of the embedding structure: t-SNE is useful for visualizing local neighborhood mixing, whereas UMAP can better preserve broader manifold organization. Agreement between the two plots was interpreted qualitatively as supporting distributional overlap; disagreement would have suggested sensitivity to the projection method. These plots are qualitative diagnostics and are interpreted together with the quantitative and downstream analyses below.

**Figure 3:**
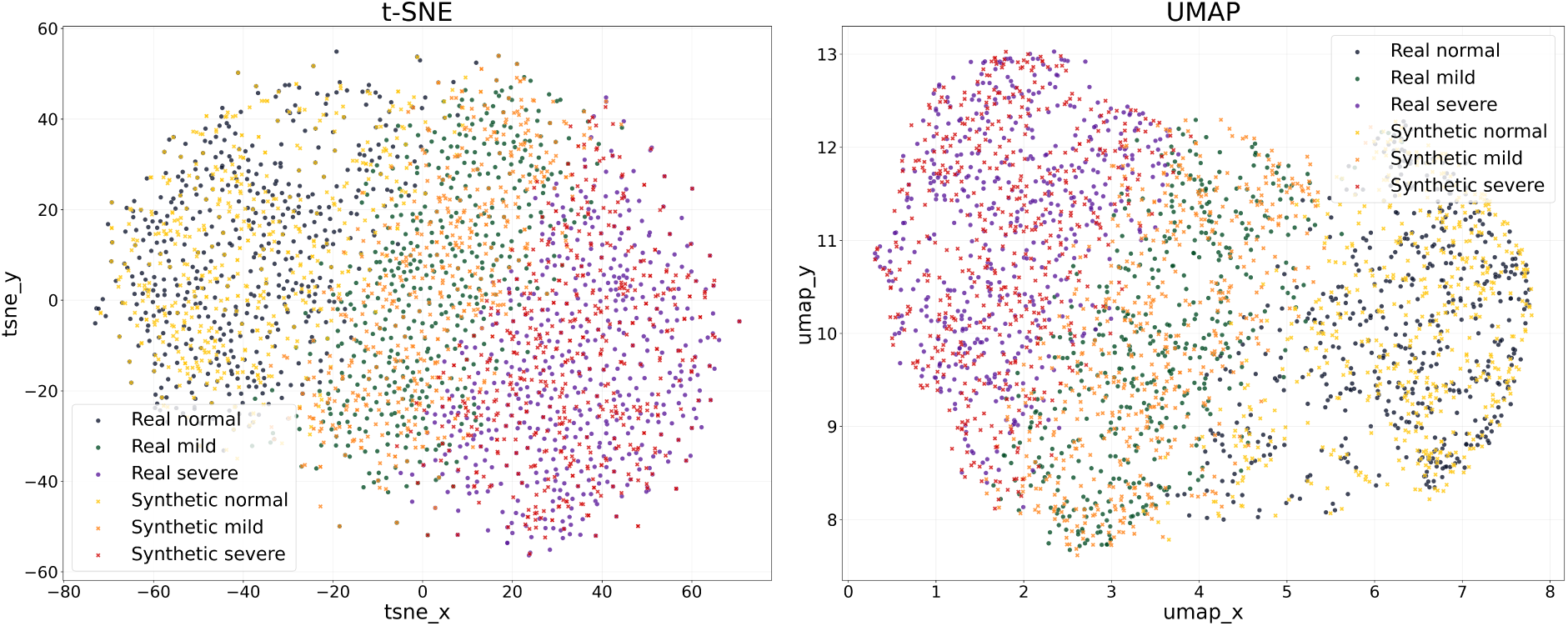
t-SNE (left) and UMAP (right) visualizations of real and synthetic echocardiographic images. Synthetic samples (yellow, orange, red) and real samples (dark blue, medium blue, purple) are shown for normal (LVEF ≥ 55%), mild dysfunction (40% ≤ LVEF < 55%), and severe dysfunction (LVEF < 40%) classes in the EchoNet-Dynamic dataset. t-SNE emphasizes local neighborhood structure, whereas UMAP provides a complementary visualization of broader manifold structure; both plots are interpreted qualitatively as checks for real–synthetic overlap.

We also examined pooled grayscale-intensity histograms as an exploratory descriptive analysis. Because no external benchmark or prespecified comparator was available, the absolute values of these histogram-based measures were not interpreted as indicating good or poor fidelity. The detailed values are provided in Supplementary Table S1. These measures summarize global grayscale-intensity profiles only; they do not evaluate spatial organization, anatomical fidelity, or the full image distributions. We considered them alongside the t-SNE and UMAP plots, discriminator analysis, expert review, and downstream classification results.

#### 3.1.1 Real-versus-Synthetic Discriminator

We evaluated real-versus-synthetic separability using the held-out discriminator described in the Methods. An AUROC of 0.5 would indicate no discrimination under this task; the observed value is interpreted together with its confidence interval and the partition design.

The discriminator achieved AUROC values of 0.907 (95% CI, 0.887–0.925) on the training set, 0.885 (95% CI, 0.853–0.914) on the validation set, and 0.612 (95% CI, 0.537–0.692) on the held-out test set. The held-out discriminator AUROC of 0.612 (95% CI, 0.537–0.692) indicated residual but limited separability between real and synthetic echocardiograms. The decrease from training and validation AUROC to held-out test AUROC indicates that discriminator performance did not generalize strongly to the test partition. Because the held-out test partition was smaller than the training partition by design, this estimate should be interpreted with its confidence interval; the train–test difference may reflect limited precision, partition-specific overfitting, or both, and does not by itself establish a mechanism.

#### 3.1.2 Expert Evaluation Demonstrates Perceptual Ambiguity but Limited Diagnostic Adequacy

We conducted a blinded expert survey to assess the perceptual and clinical fidelity of synthetic CA echocardiograms. The survey included 20 single-image evaluations, consisting of 10 real and 10 synthetic CA echocardiograms. Each image was assessed for echocardiographic view, identifiable anatomical structures, brightness, region-of-interest (ROI) presentation, diagnostic adequacy, confidence, visual realism on an ordinal scale, and perceived image source.

Completed surveys were available from 17 respondents, including 6 physicians and 11 cardiac ultrasound technicians.

Evaluators did not consistently identify synthetic images as synthetic. In the real-versus-synthetic perception question, 65.3% of synthetic-image responses were either classified as real or marked as unsure, rather than being confidently identified as synthetic. Specifically, 24.4% of synthetic responses were judged as probably or definitely real, and 40.9% were marked as unsure. This indicates perceptual ambiguity, not clinical interchangeability. Results are summarized in Table 2.

**Table 2:** Expert evaluation of real and synthetic CA echocardiograms. Percentages are computed from all non-missing single-image responses pooled across physicians and technicians (n = 199 real, n = 193 synthetic evaluations). Denominators may vary across survey items because missing responses were excluded item-wise.

| Metric | Real | Synthetic |
| --- | --- | --- |
| Brightness adequate/optimal | 60.2% | 47.4% |
| ROI presentation adequate/optimal | 57.3% | 19.2% |
| Clinically usable (Yes + Maybe) | 70.9% | 27.5% |
| Strictly diagnostic quality | 31.7% | 3.6% |
| Confidence: Agree/Strongly Agree | 78.9% | 50.5% |
| Visual realism: Agree/Strongly Agree | 65.0% | 31.1% |
| Source classification: unsure | 27.6% | 40.9% |
| Not confidently identified as synthetic | 87.9% | 65.3% |
| Avg. identifiable structures (per response) | 2.2 | 1.1 |

The survey also showed that synthetic images preserved some recognizable anatomical information. Although they were less frequently rated as fully diagnostic than real images, 27.5% of synthetic-image responses were considered clinically usable when including both “Yes — Diagnostic quality” and “Maybe — Marginal quality” responses. In addition, 47.4% of synthetic-image responses rated brightness as adequate or optimal, and 50.5% indicated agreement or strong agreement with confidence in image interpretation. Synthetic images contained identifiable anatomical information, with an average of 1.1 structures identified per response, compared with 2.2 for real images.

We further quantified response uncertainty using the normalized categorical entropy defined in the Methods. Normalized entropy was averaged within real and synthetic image groups for each question, with higher values indicating greater evaluator uncertainty or disagreement. Synthetic images showed greater evaluator uncertainty for several perceptual questions, whereas lower entropy for diagnostic adequacy and ROI presentation reflected more consistent recognition of remaining clinical limitations.

The paired-image comparison task provided a descriptive assessment of perceptual ambiguity. Across 85 paired realism responses, 40.0% were marked as “Cannot Decide”, while 31.8% preferred the real image and 28.2% preferred the synthetic image. These proportions are descriptive and do not establish a preference.

The survey showed recognizable cardiac structure in synthetic echocardiograms, but lower diagnostic adequacy than in real images. These findings do not establish downstream classification utility or clinical interchangeability.

### 3.2 Secondary Synthetic-Only Task-Retention Diagnostic

We evaluated synthetic-only task retention using two paradigms following Tian et al. [26]: Train-on-Real-Test-on-Real (TRTR), which establishes a real-only reference benchmark, and Train-on-Synthetic-Test-on-Real (TSTR), which tests synthetic-only training on held-out real transthoracic echocardiograms. This analysis was a secondary diagnostic and is distinct from the retained all-view two-step augmentation endpoint reported in Table 3.

**Table 3:** Duke two-step and EchoNet-LVH single-stage classification performance. AU-ROC and ΔAUROC values are reported with 95% confidence intervals. ΔAUROC compares real-plus-synthetic training with real-only training using the same held-out real test set. The Duke CA bootstrap baseline used resampled real minority-class training examples matched to the number of synthetic samples.

| Task | Real-only AUROC<br>(95% CI) | Bootstrap AUROC<br>(95% CI) | Real-plus-synthetic AUROC<br>(95% CI) | ΔAUROC<br>(95% CI) |
| --- | --- | --- | --- | --- |
| Duke CA, all views | 0.883 (0.831–0.927) | 0.899 (0.856–0.934) | 0.924 (0.883–0.955) | 0.041 (0.013–0.069) |
| EchoNet-LVH | 0.832 (0.784–0.874) | – | 0.864 (0.824–0.901) | 0.032 (0.021–0.044) |

Within this secondary diagnostic, the synthetic-only TSTR model retained substantial, although not complete, disease-discriminative information relative to the corresponding real-only TRTR benchmark (Figure 4). The TRTR model achieved AUROC = 0.906 (95% CI, 0.859–0.941) and AUPRC = 0.613, while the TSTR model achieved AUROC = 0.889 (95% CI, 0.842–0.936) and AUPRC = 0.575. The corresponding AUROC difference was -0.017 (95% CI, -0.075–0.036), and the TSTR AUPRC was 0.038 lower than the TRTR AUPRC. These values should not be interpreted as the real-only or real-plus-synthetic AUROC values for the two-step augmentation comparison in Table 3.

**Figure 4:**
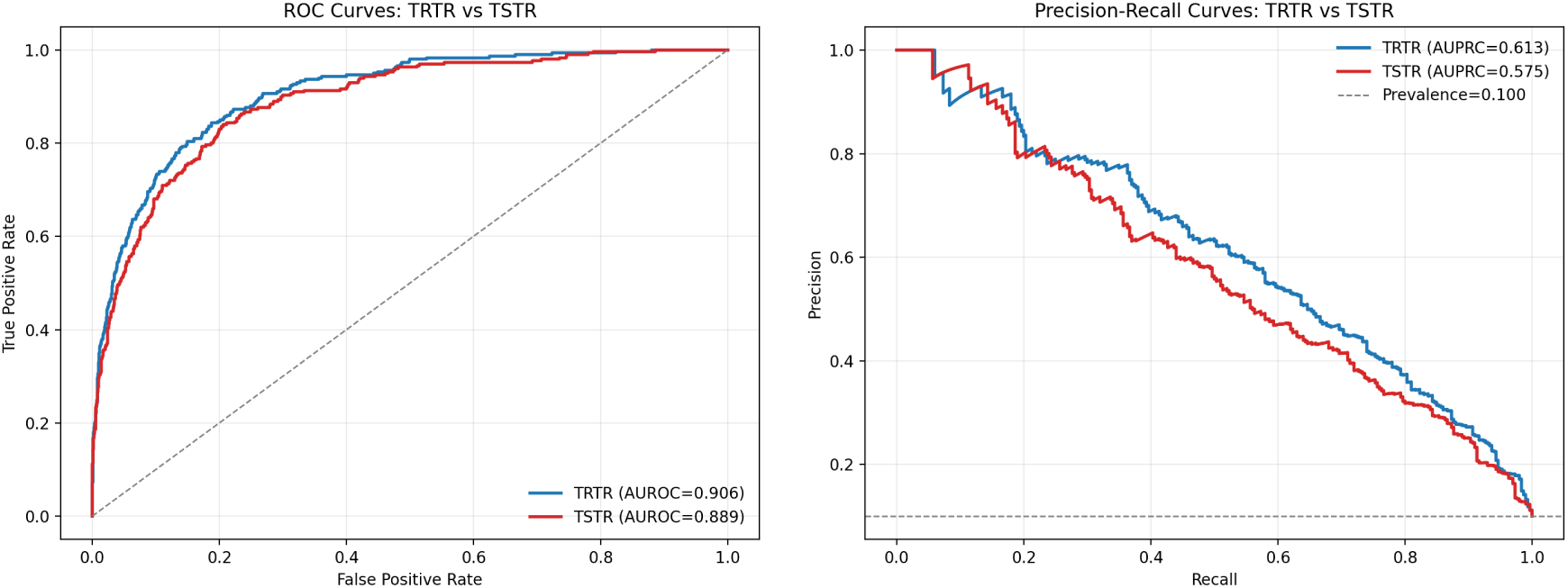
ROC and precision–recall curves for the secondary TRTR/TSTR task-retention diagnostic. The real-only TRTR model achieved AUROC = 0.906 (95% CI, 0.859– 0.941) and AUPRC = 0.613. The synthetic-only TSTR model achieved AUROC = 0.889 (95% CI, 0.842–0.936) and AUPRC = 0.575. This diagnostic is distinct from the retained all-view two-step augmentation endpoint.

### 3.3 Synthetic Augmentation in Duke and External Analyses

We evaluated synthetic augmentation using the two-step CNN framework for the Duke CA task and a single-stage binary CNN for EchoNet-LVH. The Duke analysis used all available echocardiographic views rather than automated view-classified subsets. For each task, we report the AUROC without synthetic augmentation, the AUROC with synthetic augmentation (TRSTR), and the paired AUROC difference (ΔAUROC). For the Duke CA analysis, the bootstrap baseline is included as a class-balance comparison.

In the Duke all-view CA analysis, adding synthetic CA images produced a higher two-step CNN AUROC point estimate, from 0.883 (95% CI, 0.831–0.927) to 0.924 (95% CI, 0.883–0.955), corresponding to ΔAUROC = 0.041 (95% CI, 0.013–0.069; Table 3). Bootstrap resampling yielded an intermediate AUROC of 0.899 (95% CI, 0.856–0.934). The comparison suggests that the observed point-estimate difference may reflect more than ordinary minority-class resampling alone, but it does not establish a mechanism or a statistically confirmed benefit beyond this comparator.

EchoNet-LVH was evaluated using the video partitions, wall-thickness phenotype definition, and synthetic-training protocol described in the Methods. Synthetic samples were generated using only the training data, and the validation and test sets contained exclusively real echocardiograms. In this external wall-thickness task, the single-stage CNN AUROC increased from 0.832 (95% CI, 0.784–0.874) to 0.864 (95% CI, 0.824–0.901), corresponding to ΔAUROC = 0.032 (95% CI, 0.021– 0.044).

## 4 Discussion

Across the evaluated echocardiographic tasks, synthetic augmentation was associated with higher held-out AUROC in the all-view Duke CA analysis and the single-stage EchoNet-LVH wall-thickness analysis. The results suggest that diffusion-generated echocardiograms may be useful for selected training tasks, but they are not a replacement for real data.

The Duke CA experiment demonstrates the use case most directly. CA combines an uncommon target class with phenotypic overlap with LVH, creating a setting in which additional minority-class examples may be useful. The two-step design separates detection of structural abnormality from differentiation of CA and LVH. A secondary TRTR/TSTR diagnostic suggested that synthetic-only training retained substantial, although incomplete, task-relevant information, while real-plus-synthetic training produced higher AUROC than real-only training in the all-view Duke two-step analysis. The bootstrap comparison suggests that the observed difference may reflect more than class rebalancing alone, but it does not establish a mechanism.

The external analyses addressed different questions. EchoNet-Dynamic was used for LVEF-conditioned generation and external fidelity evaluation, not for CA detection or multi-view validation. EchoNet-LVH evaluated wall-thickness phenotype classification with a single-stage binary CNN in a standardized external cohort and showed a positive AUROC difference after synthetic augmentation.

The fidelity analyses also measure distinct properties. The pooled grayscale histogram statistics summarize global intensity profiles and do not assess spatial organization or anatomical fidelity. The held-out discriminator AUROC of 0.612 indicates residual but limited separability between real and synthetic images. Expert responses showed perceptual ambiguity, but synthetic images had lower diagnostic adequacy and lower strict diagnostic-quality ratings than real images. Thus, perceptual ambiguity, histogram similarity, and downstream augmentation utility should not be interpreted as evidence of clinical interchangeability.

The study has several limitations. The primary cohort was collected at a single academic medical center, and performance in other institutions, scanner environments, and patient populations remains uncertain. CA examinations spanned June 2004–March 2024, whereas the control cohorts were collected primarily during January 2015–December 2023; acquisition era, scanner, and protocol may therefore be correlated with phenotype. The current analysis did not include a contemporaneous-period or scanner-matched sensitivity analysis. CA labels were based on expert-adjudicated clinical diagnoses, and the manuscript does not provide complete subtype-specific reference-standard information or a quantitative LVH threshold sufficient to exclude occult CA from every LVH control. The reported cohort table provides encounter counts, but detailed age, sex, scanner/vendor, acquisition-year distributions, and images-per-encounter summaries were not available in the current analysis. The encounter-level split also does not establish that repeated encounters from the same patient were confined to one partition; patient-level leakage therefore cannot be excluded without an additional patient-identity audit. The study used the first frame from each retained video and therefore did not model temporal information; in EchoNet-LVH, this first-frame input was not necessarily the end-diastolic measurement frame. Images were generated and evaluated at 112 × 112 pixels, lower than typical clinical acquisition resolution. The classification tasks did not incorporate multimodal clinical variables. External evaluation was limited in view coverage. In addition, generation and augmentation procedures were task-specific, and formal privacy, memorization, and nearest-neighbor analyses were not performed.

Stage-specific performance for the Duke two-step model and a full baseline comparison beyond bootstrap resampling were not included in the current analysis. The primary augmentation results therefore should not be interpreted as establishing superiority over cost-sensitive learning, balanced sampling, focal loss, or other conventional augmentation strategies.

Future work should evaluate video diffusion models, higher-resolution synthesis, and additional standardized echocardiographic views. Structural and functional fidelity measures should complement global intensity statistics, and diversity, nearest-neighbor, and memorization analyses should be reported. Multi-site evaluation, more stable external splits, formal privacy assessment, and class-conditional anatomical measurements may help determine when synthetic echocardiograms provide reliable training information.

## Data availability

The Duke CA dataset contains protected health information and is not publicly available owing to patient privacy restrictions. De-identified summary data and the trained diffusion model weights will be made available upon reasonable request to the corresponding author (A.R.Z.), subject to institutional data-use agreement approval. EchoNet-Dynamic and EchoNet-LVH were accessed under their respective data-use agreements and public-data access procedures [2, 27].

## Code availability

The code used in this study, including the analysis code and trained model scripts, is available on GitHub at https://github.com/Lyu-Pengfei/echosynth.

## Consent to participate

The requirement for informed consent was waived by the Duke University Health System Institutional Review Board due to the retrospective nature of the study.

## Funding and grants

The research is supported in part by NIH Grant R01HL168940. Sreekanth Vemulapalli reports grants or contracts from the American College of Cardiology, American Heart Association, U.S. Food and Drug Administration, National Institutes of Health (R01 and UG3/UH3), Edwards Life-sciences, Abbott Vascular, Kardigan Bio, and Idorsia.

## Competing interests

Sreekanth Vemulapalli reports consulting or advisory relationships with Edwards Lifesciences, Abbott Vascular, Medtronic, and Eli Lilly. The other authors declare no competing interests.

## Supplementary Information

### A Supplementary Fidelity Metrics

**Table S1:**
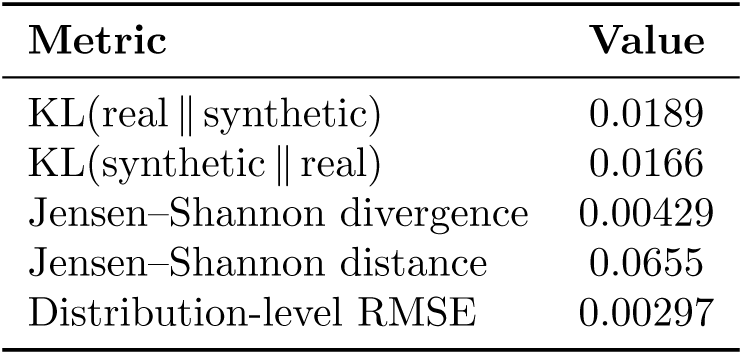
Supplementary exploratory histogram-based diagnostics. All quantities were calculated from the same pooled 256-bin grayscale-intensity histograms. No external benchmark or prespecified comparator was available; therefore, these values are reported descriptively and are not interpreted as evidence of good or poor fidelity or as independent fidelity analyses.

### B Expert Survey Questionnaire

#### Purpose and Image Selection

This questionnaire is part of a research study investigating the visual realism, perceived source, and clinical adequacy of diffusion-model-generated synthetic echocardiographic images relative to real clinical images.

In the questionnaire, the participants will be asked to evaluate a set of echocardiographic images drawn from two sources: (1) de-identified real echocardiograms acquired at Duke University Health System (DUHS), and (2) synthetic echocardiograms generated by a class-conditional diffusion model trained on the same patient cohort.

Images were randomly selected and shuffled; neither the participant nor the survey platform retains information about which images are real or synthetic until after all responses are collected.

##### No patient-identifiable information is included

All real images have been de-identified in accordance with HIPAA and the study protocol approved by the Duke University Institutional Review Board (IRB). Participation is voluntary, all responses are anonymous, and data will be reported only in aggregate.

The identical questionnaire was distributed to two independent groups: physicians (MDs) and echocardiography technicians, such that responses can be analyzed separately by professional role and compared across groups.

#### Response format

◯ indicates select one option only; □ indicates select all that apply.

> **Echocardiographic Image Evaluation for MDs / Technicians**

#### Welcome page

Welcome to the Echocardiographic Image Evaluation Survey for MDs / Technicians. You will be shown a series of echocardiographic images. Some are real, others are synthetic. Your task is to evaluate each image as you would in clinical practice. For each image, please answer a few questions about its diagnostic utility, visual realism, and perceived source. The survey should take approximately 10–15 minutes to complete. All responses are anonymous and will be used for research purposes only. Click “Next” to begin.

#### Section 1: Individual Image Evaluation

You will be shown one echocardiographic image at a time and asked to answer the following questions based on each image. For each image, please answer the following questions based on what you observe.

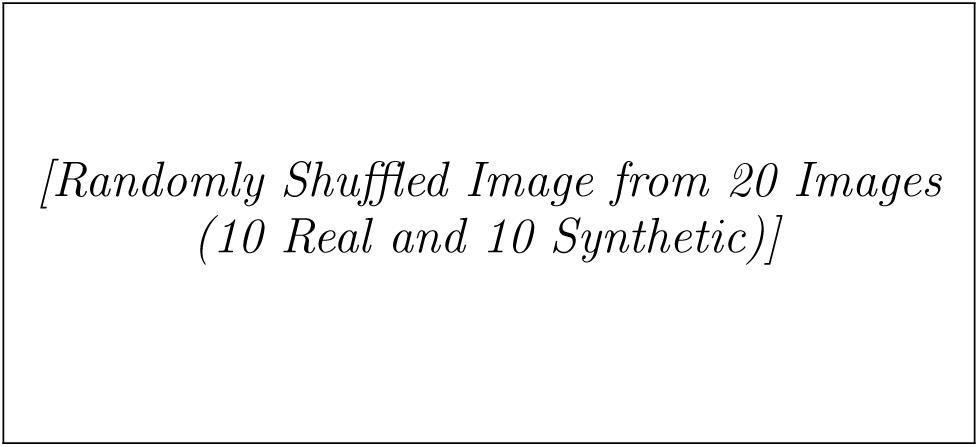

1. Which echocardiographic view is shown in this image?

- Parasternal Long-Axis
- Parasternal Short-Axis
- Apical 4-Chamber
- Apical 2-Chamber
- Subcostal
- Suprasternal
- ther / Not Sure
2. Which structures can you clearly identify in this image? (Select all that apply)

□ Left Ventricle
□ Right Ventricle
□ Left Atrium
□ Right Atrium
□ Mitral Valve
□ Aortic Valve
□ Interventricular Septum
□ LV Apex
□ Aorta
□ IVC
□ None / Not Sure
3. Is the brightness level appropriate?

- Optimal
- Adequate
- Inadequate
4. Is the Region of Interest presented well?

- Optimal
- Adequate
- Inadequate
5. Would you consider this image adequate for diagnostic interpretation?

- Yes — Diagnostic quality
- Maybe — Marginal quality
- No — Not diagnostic
6. Please rate your agreement with the following statements:

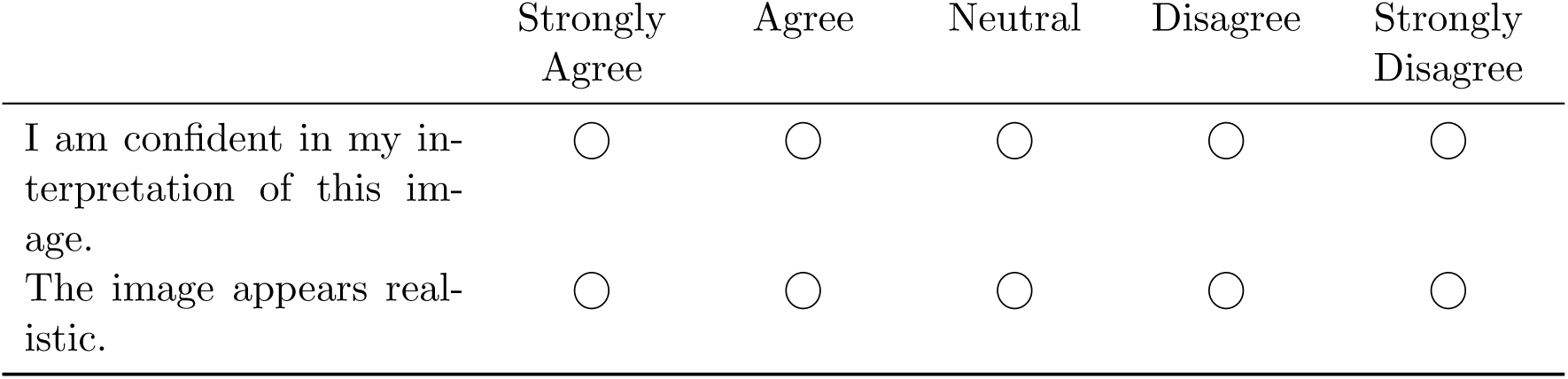
7. In your opinion, this image is:

- Definitely Real
- Probably Real
- Unsure
- Probably Synthetic
- Definitely Synthetic

#### Section 2: Pairwise Image Comparison

You will be shown echocardiographic images in pairs. Each image within a pair may be real or synthetic. Please answer the following questions for each pair.

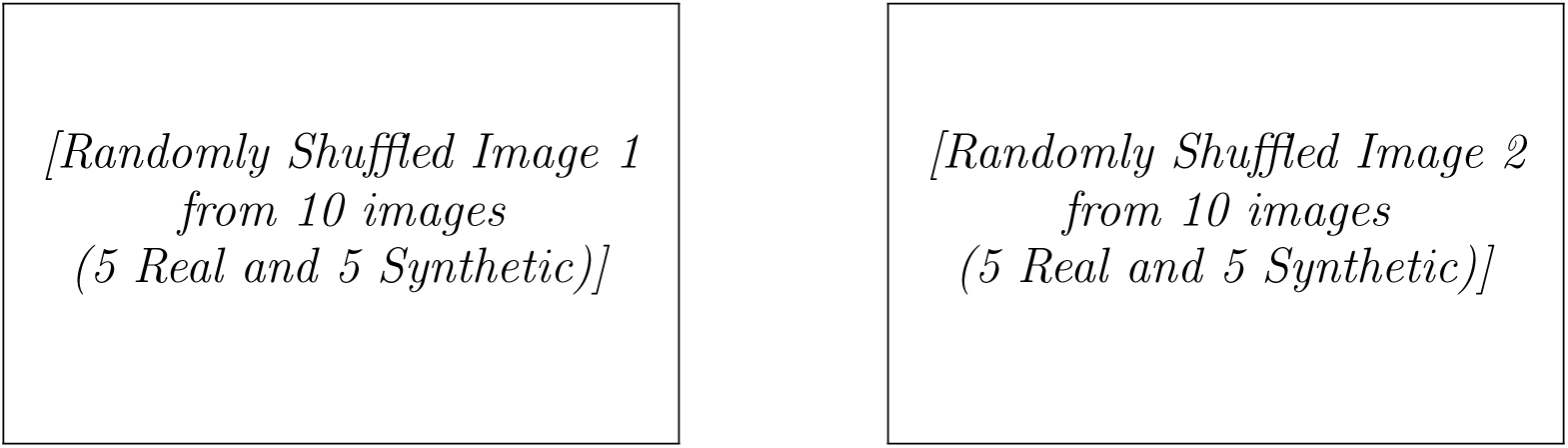

1. Which image appears more realistic? o The First Image

- The Second Image
- Cannot Decide
2. Which image would you prefer to use for clinical diagnosis?

- The First Image
- The Second Image
- Neither — Both are inadequate
- Both are equally acceptable

#### Final Comments (Optional)

Do you have any comments about the images or the survey design?

## Notes

### Author Declarations

The Duke University Health System Institutional Review Board gave ethical approval for this work and waived the requirement for informed consent due to the retrospective nature of the study.

